# KDClassifier: Urinary Proteomic Spectra Analysis Based on Machine Learning for Classification of Kidney Diseases

**DOI:** 10.1101/2020.12.01.20242198

**Authors:** Wanjun Zhao, Yong Zhang, Xinming Li, Yonghong Mao, Changwei Wu, Lijun Zhao, Fang Liu, Jingqiang Zhu, Jingqiu Cheng, Hao Yang, Guisen Li

## Abstract

**Background:** By extracting the spectrum features from urinary proteomics based on an advanced mass spectrometer and machine learning algorithms, more accurate reporting results can be achieved for disease classification. We attempted to establish a novel diagnosis model of kidney diseases by combining machine learning with an extreme gradient boosting (XGBoost) algorithm with complete mass spectrum information from the urinary proteomics.

**Methods:** We enrolled 134 patients (including those with IgA nephropathy, membranous nephropathy, and diabetic kidney disease) and 68 healthy participants as a control, and for training and validation of the diagnostic model, applied a total of 610,102 mass spectra from their urinary proteomics produced using high-resolution mass spectrometry. We divided the mass spectrum data into a training dataset (80%) and a validation dataset (20%). The training dataset was directly used to create a diagnosis model using XGBoost, random forest (RF), a support vector machine (SVM), and artificial neural networks (ANNs). The diagnostic accuracy was evaluated using a confusion matrix. We also constructed the receiver operating-characteristic, Lorenz, and gain curves to evaluate the diagnosis model.

**Results:** Compared with RF, the SVM, and ANNs, the modified XGBoost model, called a Kidney Disease Classifier (KDClassifier), showed the best performance. The accuracy of the diagnostic XGBoost model was 96.03% (CI = 95.17%-96.77%; Kapa = 0.943; McNemar’s Test, *P* value = 0.00027). The area under the curve of the XGBoost model was 0.952 (CI = 0.9307-0.9733). The Kolmogorov-Smirnov (KS) value of the Lorenz curve was 0.8514. The Lorenz and gain curves showed the strong robustness of the developed model.

**Conclusions:** This study presents the first XGBoost diagnosis model, i.e., the KDClassifier, combined with complete mass spectrum information from the urinary proteomics for distinguishing different kidney diseases. KDClassifier achieves a high accuracy and robustness, providing a potential tool for the classification of all types of kidney diseases.

## INTRODUCTION

Chronic kidney disease (CKD), as a major public health problem, has more than a 10% global incidence rate and is a significant burden globally^1, 2^. Persisting renal damage and loss of renal function are the main clinical characteristics of CKD. Despite the continuous effort of nephropathologists, the incidence, prevalence, mortality rate, and disability-adjusted life years of CKD remain extremely high, and have even increased significantly in recent decades^2^. Kidney diseases are mainly evaluated based on persistent proteinuria, hematuria, and clinical impairment of the renal function, as well as a decrease in the glomerular filtration rate (GFR)^3, 4^. However, the clinical characteristics of kidney diseases with different pathological categories are obviously different, including primary glomerular diseases such as IgA nephropathy (IgAN) and membranous nephropathy (MN), and secondary glomerular diseases such as diabetic kidney disease (DKD). How to distinguish the diseases more easily and early, and how to treat them more precisely, are important aspects for improving the outcomes of CKD.

With the innovation of puncture biopsy technology, renal biopsies have become the most critical technology for the pathological diagnosis of kidney diseases in recent years as well for the elucidation of various renal diseases^5-7^. A renal biopsy is the gold standard in terms of diagnosis, treatment, and prognosis of kidney disease through a pathologic analysis. A series of important advances in renal pathology have promoted our understanding of the pathogenesis of renal diseases, and in the future, an artificial intelligence assisted pathological analysis will expand our understanding of renal pathological lesions and the pathogenesis of kidney disease^7-9^. However, as an invasive procedure, a kidney biopsy may incur some ineluctable complications, the most frequent being macrohematuria with or without the need for a blood transfusion^10, 11^. In addition, many patients are unable to receive a renal biopsy because of relative or absolute contraindications. It is therefore necessary to find novel noninvasive biomarkers or methods to improve the diagnostic efficiency, monitoring, and treatment of CKD.

Some existing studies have shown that urine, serum metabolite, and protein have potential clinical application as biomarkers^12-14^. Proteins are considered as the final products of gene-environment interactions and a physiological steady-state. A single highly specific and unique biomarker (such as an M-type phospholipase A2 receptor for MN) is certainly the best choice^15^; however, such biomarkers are unavailable for clinically noninvasive diagnosis in numerous kidney diseases, such as IgAN or DKD. The measurement of various urinary proteins can be combined with the available clinical biochemistry indexes, which have the potential for clinical diagnosis, patient stratification, and therapeutic monitoring^16^. Proteomics provide new insight into biomarker discovery and have dramatically widened our appreciation of pathological mechanisms. New analytical tools with high accuracy have made proteomics easier and quantifiable, allowing an exploration of information from biological samples^17^.

The mass spectra of urinary proteome produced by liquid chromatography tandem mass spectrometry (LC-MS/MS) are big datasets containing rich information. Existing software cannot interpret all spectral information. With the development of mass spectrometry and machine learning algorithms, the extraction of spectrum features from the urinary proteome of each disease entity based on an advanced mass spectrometer and machine learning algorithms can save a lot of time and obtain more accurate reporting results. Therefore, we believe that the use of all mass spectrum information from a urinary proteome, as provided through advanced mass spectrometry, can be an effective potential research direction to improve the accuracy of CKD diagnosis.

With this study, we attempted to train and validate a diagnostic machine learning model using more than 600,000 mass spectra from the urinary proteome produced by LC-MS/MS in CKD patients. This method permits the rapid extraction of the spectrum features in human urine (including soluble proteins, exosomes, and other membrane elements). We compared four machine learning models, namely, an artificial neural network (ANN), a support vector machine (SVM), a decision tree (DT), and extreme gradient boosting (XGBoost). We chose the most accurate model and evaluated its performance in terms of the classification of CKD patients and a group of healthy control (HC) participants. Finally, the XGBoost model, called a Kidney Disease Classifier (KDClassifer), showed the best performance in distinguishing different CKD patients using the mass spectra from the urinary proteome of the IgAN, MN, and DKD patients, and the HC group. The mass spectra data on the urinary proteomics were deposited into the ProteomeXchange Consortium through the PRIDE partner repository using the dataset identifier **PXD018996**.

## RESULTS

### Basic Characteristics of Kidney Disease Dataset

In this study, we enrolled 134 CKD patients with different pathological classifications (IgAN = 50, MN = 50, and DKD = 34) and 68 healthy control participants (HC = 68), the characteristics of which are shown in Table 1. Among the four groups, the gender ratio of each group was between 0.5 and 2. The average ages of the four groups were ranked from oldest to youngest as DKD, MN, HC, and IgAN. The difference in average age was statistically significant (*P* < 0.01, ANOVA test). However, this was consistent with the age distribution trend of different types of kidney disease. The workflow of this study is shown in Figure 1. The urinary proteome was treated using an ultrafiltration tube-assisted digestion method that can maintain the urinary exosomes and other membrane elements. The tryptic peptides were then analyzed using a high-resolution mass spectrometer. Finally, a total of 610,102 urinary proteomic mass spectra were produced for training and validation of the diagnostic model, including 165,521 spectra from the IgAN group, 151,159 spectra from the MN group, 46,187 spectra from the DKD group, and 247,235 spectra from the HC group. All spectra in each group were randomly divided into a training dataset (80%) and a validation dataset (20%). As shown in Figure 2, the distribution of the different patient types in the training and validation datasets and the proportion of kidney disease types were nearly consistent.

**Table 1.**
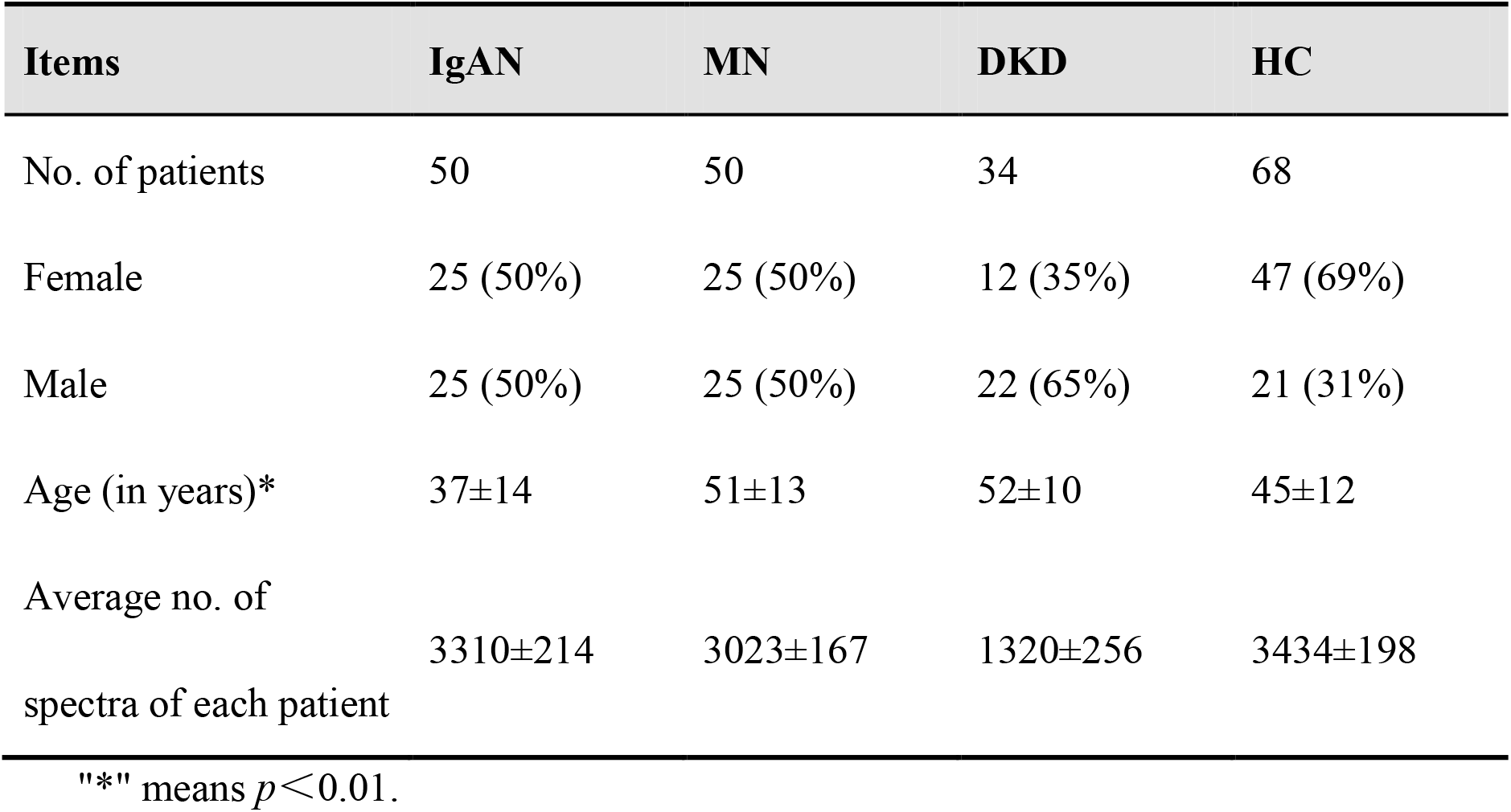
Basic information of the patients and healthy control group

| Items | IgAN | MN | DKD | HC |
| --- | --- | --- | --- | --- |
| No. of patients | 50 | 50 | 34 | 68 |
| Female | 25 (50%) | 25 (50%) | 12 (35%) | 47 (69%) |
| Male | 25 (50%) | 25 (50%) | 22 (65%) | 21 (31%) |
| Age (in years)* | 37±14 | 51±13 | 52±10 | 45±12 |
| Average no. of spectra of each patient | 3310±214 | 3023±167 | 1320±256 | 3434±198 |

"" means $p < 0.01$ .

**Figure 1.**
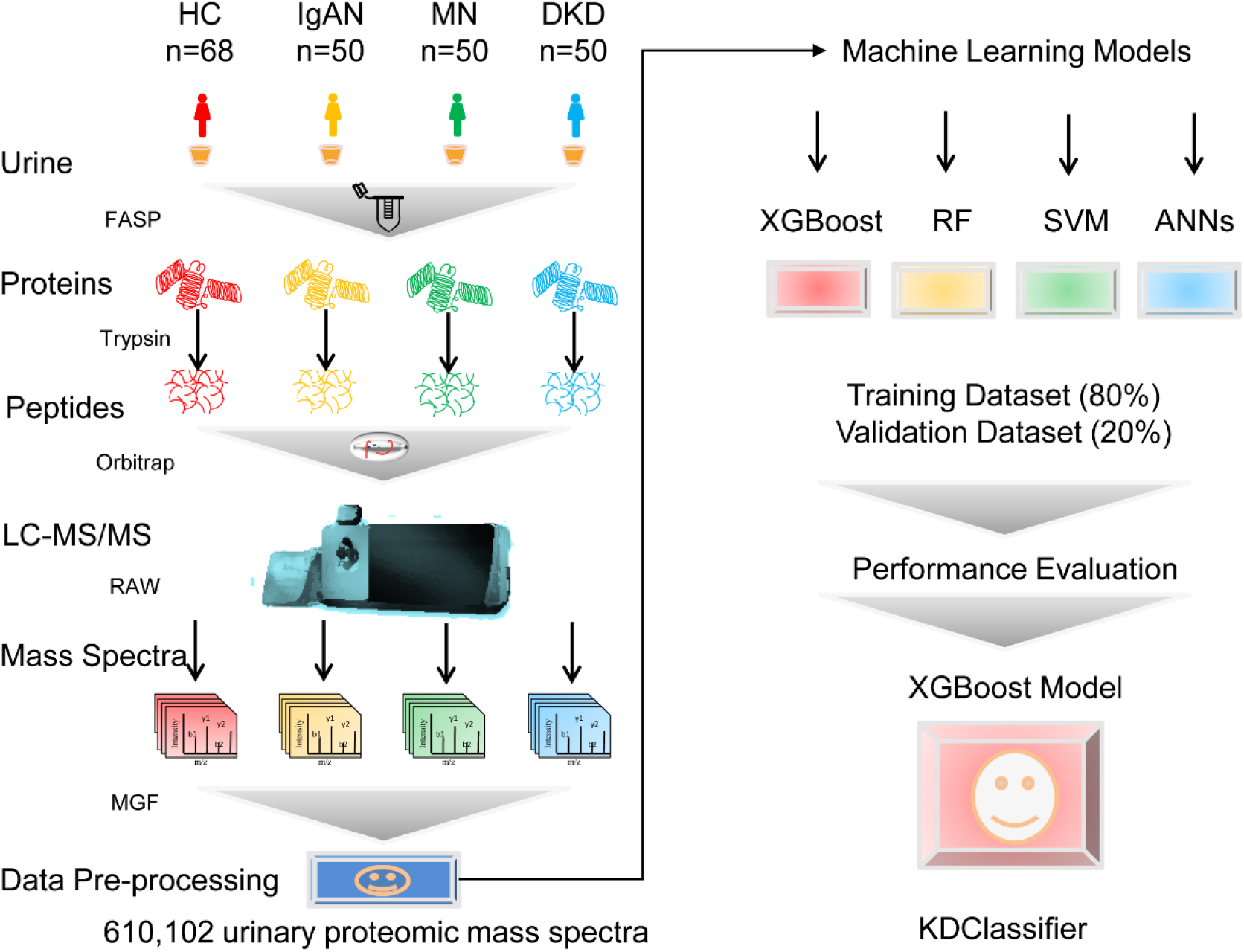
Workflow of spectrum analysis from urinary proteomics based on machine learning for classification of kidney diseases.

**Figure 2.**
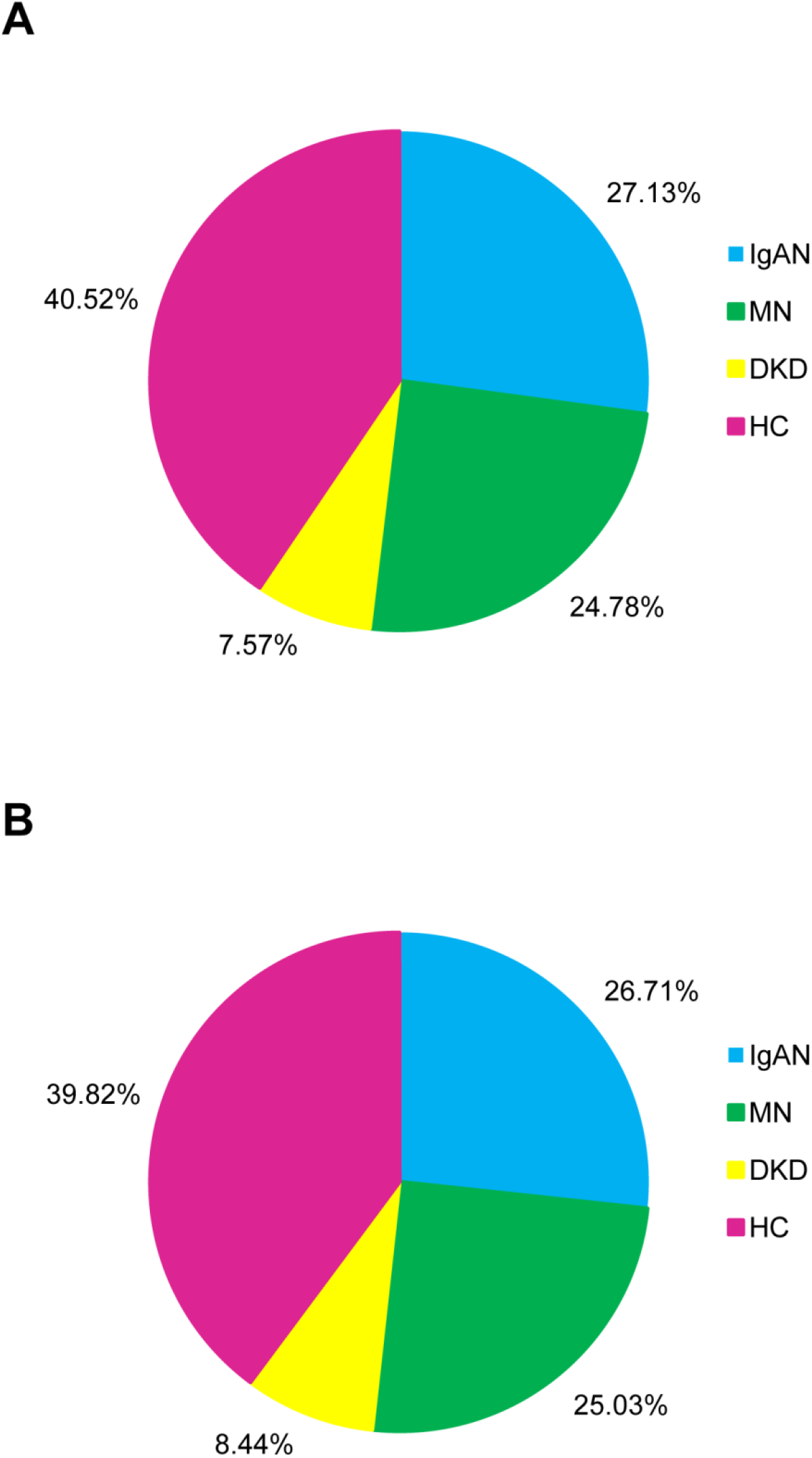
Proportion of three types of CKD and healthy control samples for (A) training and (B) validation of the XGBoost model.

### Comparison of Diagnostic Accuracy between XGBoost and other Machine Learning Models

After training, the accuracy of the diagnostic XGBoost model was validated as 96.03% (CI = 95.17%–96.77%) (Table 2). The Kapa value was 0.943 and the P-value of the McNemar’s Test was 0.00027, which showed the perfect performance of XGBoost. The RF, SVM, and ANN models were trained in the same way, with an accuracy of 92.35%, 86.12%, and 87.28%, respectively. The accuracy of all machine learning models tested was relatively high. However, compared with the other models, XGBoost achieved the best performance, and was thus applied as our machine learning algorithm (Table 2).

**Table 2.** Accuracy of different models in training and validation datasets

| Model | Accuracy (CI 95%) |  |
| --- | --- | --- |
|  | Training dataset | Validation dataset |
| Random Forest | 96.36% | 92.35% |
|  | (95.63%~97.18%) | (91.28%~94.23%) |
| Support Vector Machine | 92.67% | 86.12% |
|  | (89.56%~93.43%) | (84.28%~89.71%) |
| Artificial Neural Networks | 95.12% | 87.28% |
|  | (93.96%~96.71%) | (84.27%~90.16%) |
| XGBoost | 99.21% | 96.03% |
|  | (98.89%~99.48%) | (95.17%~96.77%) |

### Classification Performance of Kidney Disease Diagnostic XGBoost Model

To characterize the performance of the diagnostic XGBoost model for different types of kidney diseases, we compared the predictive ability of this model for the three types of kidney diseases detailed above and the HC. We chose 20% of the total dataset to test. Although the number of test errors was large, the error rate was low. As shown in Table 3 and Figure 3, the false rate of the four different types (IgAN, MN, DKD, and HC) was 2.76%, 5.73%, 10.19%, and 2.37%, respectively. The XGBoost model achieved the highest error rate for DKD and the lowest error rate for IgAN. The accuracy of the three types of kidney disease and the HC were 97.67%, 96.64%, 94.86%, and 97.35%, respectively (Table 4). Although the accuracy of the diagnosis for each of the four different types was extremely high, the diagnostic accuracy of the DKD was the lowest. Specifically, comparing four performance items, namely, the sensitivity, specificity, positive predictive value, and negative predictive value, we found that the positive predictive rates for the IgAN and HC groups were relatively low, as was the sensitivity for both the MN and DKD groups. In addition, we specifically analyzed the misclassification of the four types. As shown in Figure 4, the IgAN and MN groups were relatively easy to be misjudged as the HC group, whereas the DKD and HC groups were relatively easy to be misjudged as the IgAN group.

**Table 3.**
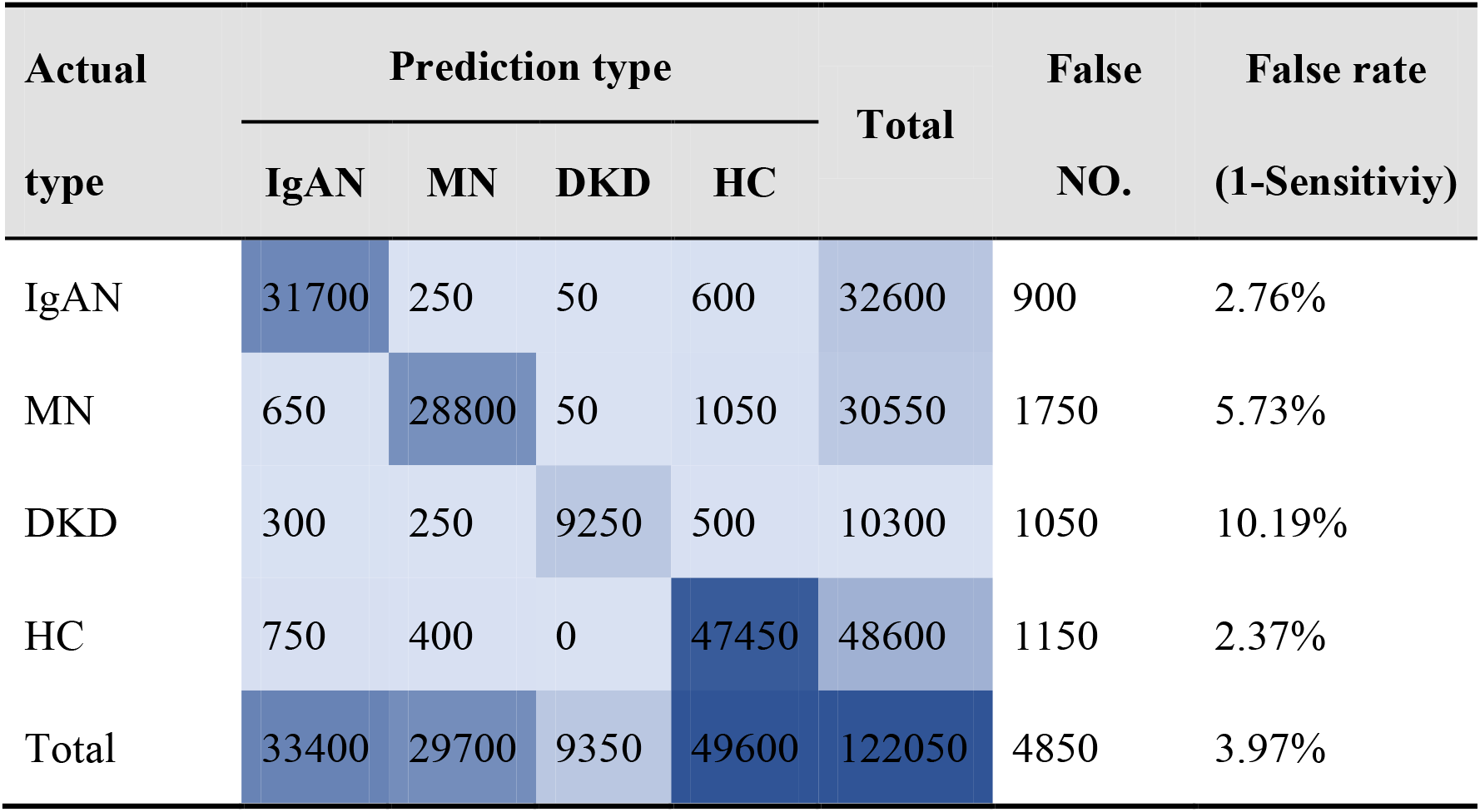
Confusion matrix of XGBoost for diagnosis of chronic kidney diseases

**Table 4.** Performance of XGBoost model for diagnosis of chronic kidney diseases

| Items | IgAN | MN | DKD | HC |
| --- | --- | --- | --- | --- |
| Sensitivity | 97.24% | 94.27% | 89.81% | 97.63% |
| Specificity | 98.10% | 99.02% | 99.91% | 97.07% |
| Pos-Pred-Value | 94.91% | 96.97% | 98.93% | 95.67% |
| Neg-Pred-Value | 98.98% | 98.11% | 99.01% | 98.41% |
| Balanced accuracy | 97.67% | 96.64% | 94.86% | 97.35% |

**Figure 3.**
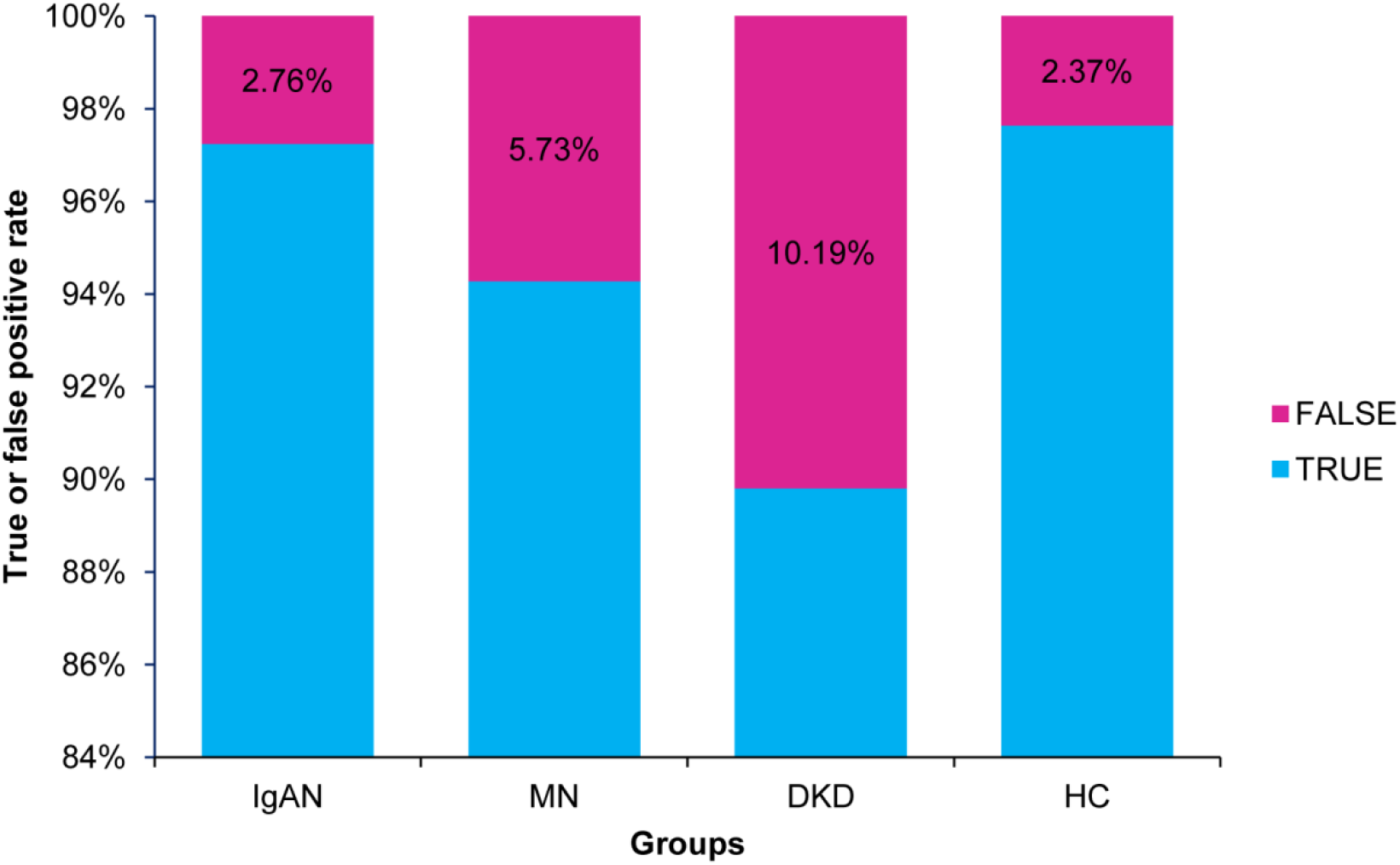
Bar chart of the diagnosis error rate of three types of CKD patients and healthy control group for validation dataset of the XGBoost model.

**Figure 4.**
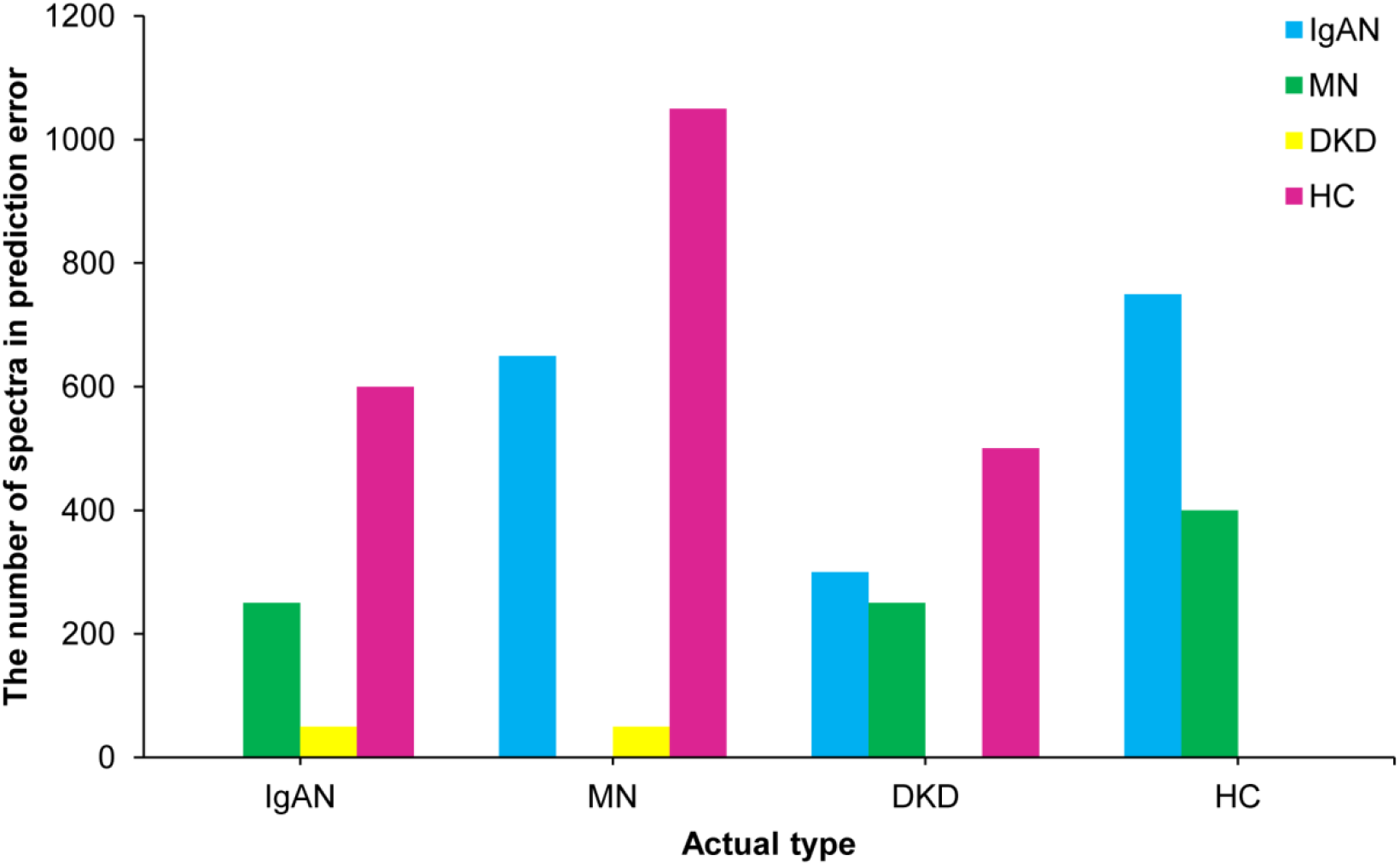
Bar chart of misclassification for three types of CKD patients and healthy control group.

### Evaluation of Diagnostic XGBoost Model for Kidney Disease

The discrimination of kidney disease when applying the diagnostic XGBoost model was assessed based on the receiver operating characteristic (ROC) curve and area under the curve (AUC) (Figure 5). The AUC value of this model was 0.952 (CI = 0.9307–0.9733), demonstrating a strong generalization. In addition, the slope of the gain curve was adequately steep. When the test sample rate was 18.7%, the TPR reached 92.3%, which showed the high TPR of the model (Figure 6). The Kolmogorov-Smirnov (KS) value of the Lorenz curve was 0.8514, which was much higher than 0.2 (Figure 7). The gain and Lorenz curves also demonstrate the strong robustness of the model.

**Figure 5.**
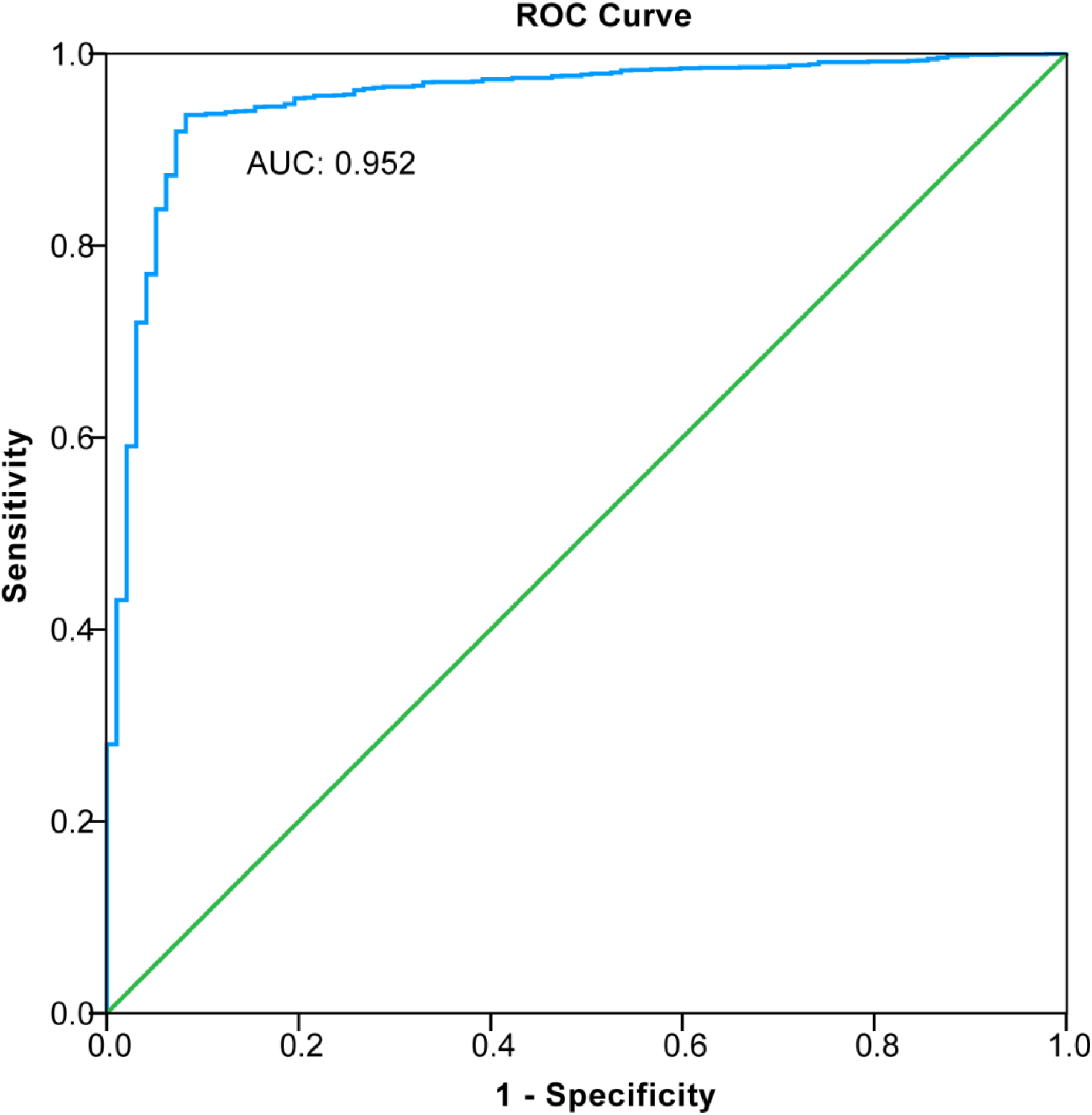
Receiver operating curve (ROC) for estimating the discrimination of XGBoost.

**Figure 6.**
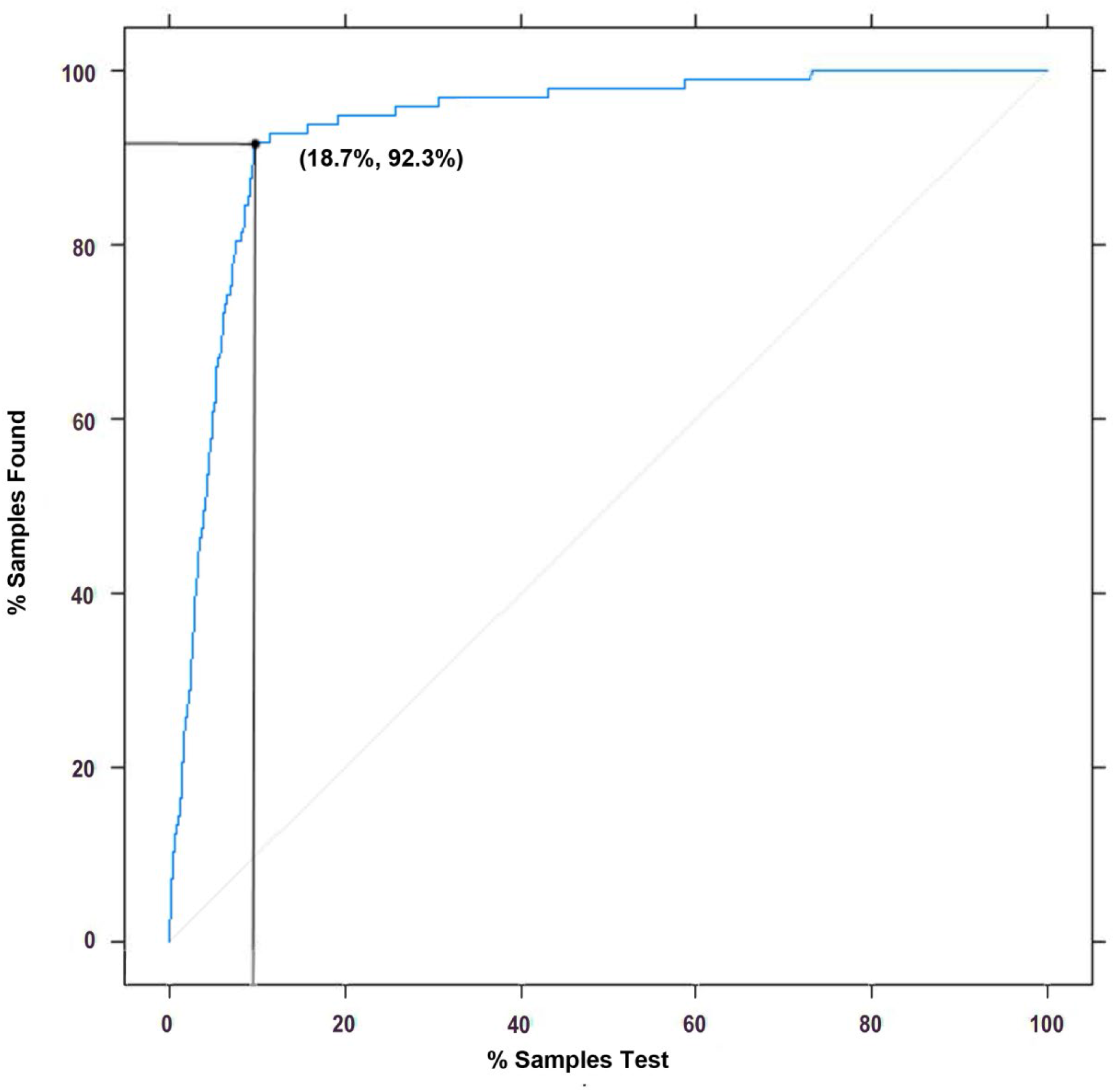
Gain plot for evaluating the overall diagnostic accuracy of the XGBoost model.

**Figure 7.**
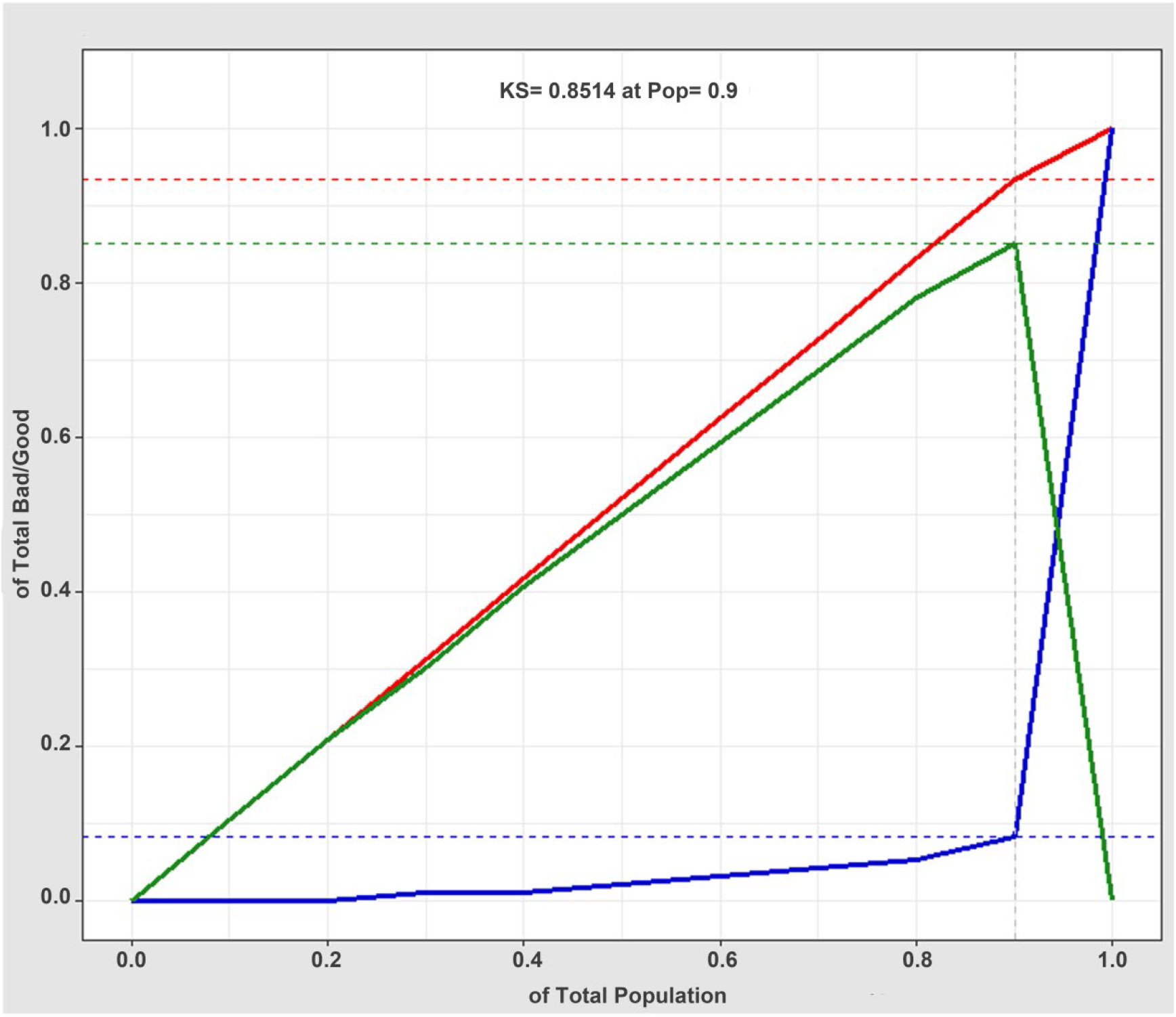
Lorenz curve (KS curve) for evaluating the goodness of fit of the XGBoost model diagnosis. The red, blue, and green lines are the true positive prediction rate, the false positive prediction rate, and the distance between the true positive prediction rate and the false positive prediction rate, respectively. The Lorenz value is the threshold value corresponding to the farthest distance between the red and blue lines, which is the threshold value that can best divide the model.

## DISSCUSSION

CKD represents a major public health issue in terms of its substantial financial burden and consumption of healthcare resources^1^. In addition, CKD is a risk factor for hypertension and cardiovascular disease, which together constitute a substantial cause of death in most societies^18^. How to accurately identify and achieve an early screening of CKD in the population has long been an important topic. The development of a non-invasive and accurate early diagnosis is needed. The diagnostic ability of a single biomarker is slightly weak, and a renal biopsy is invasive with a risk of major bleeding. With the development of mass spectrometry, we can detect the urinary proteome both quantitatively and qualitatively^17^. Our study was devoted to artificial intelligence assisted noninvasive diagnostic methods of different types of kidney disease based on mass spectra information from the urinary proteomics.

Previous studies have shown that the application of a single protein in the clinical diagnosis of CKD does not take advantage of the overall value and macro efficiency of the proteomics^17^. In addition, the feasibility of using a single protein in the clinical diagnosis of CKD requires further research and validation. The use of several or even dozens of protein panels can improve the accuracy of the diagnosis. Existing mass spectrometry applied to proteomics is used in the identification of differential proteins and the selection of individual proteins for further differential studies. In fact, the overall data from a mass spectrometry analysis is not applied. Moreover, the efficacy of its clinical application requires further evaluation. Using big data, we can apply machine learning by integrating all information from a mass spectrometry analysis. We strived to analyze all data to take full advantage of the overall efficiency of proteomic mass spectrometry. Therefore, for CKD classification, considering the comprehensiveness of a mass spectrum analysis, as the feature data of our AI algorithm, we apply a first-order mass spectrum analysis of the proteomics without further processing. Artificial intelligence algorithms such as an ANN, an SVC, a DT, and XGBoost incorporated with medical or biological experience have obtained remarkable results^19, 20^. Through the training of big datasets, a machine learning model can predict a classification. Machine learning outperforms conventional statistical methods with its ability to better identify variables, achieves a better predictive performance and a better modeling of complex relationships, has the ability to learn from multiple modules of data, and it robust to data noise. It has therefore been applied to the diagnosis of certain diseases, such as lung cancer^21^, cardiovascular disease^22^, and chronic kidney disease^23^. Machine and deep learning algorithms can not only impute missing data in the training sets they can also identify existing characteristics that we otherwise cannot recognize. Most existing machine models for CKD are based on records and detection indicators that are currently used in clinical practice^24^. However, the training data types of these models vary, and the accuracy of an artificial collection is relatively low with a poor clinical application. To date, there have been no studies on machine learning models for diagnosis based on the full spectra of CKD urinary proteomics. In addition, in many existing machine learning models, XGBoost achieves an outstanding classification performance without a high computation time and is a practical approach. XGBoost is a type of tree-structured model, the basic idea of which is to design an ensemble approach for several rule-based binary trees. GXBoost is derived from the most famous tree ensemble method, called a gradient boosting decision tree (GBDT). GXBoost has gained popularity by winning numerous machine learning competitions since its initial development^25^. Advances in big data and artificial intelligence have enabled clinicians to process information more efficiently, and make diagnosis and treatment decisions more accurately^26^. It is unquestionable that big data and artificial intelligence are transforming medicine from various perspectives, including precision medicine and clinical intelligence. Based on the big data applied in urinary proteomic mass spectra, the strategy of artificial intelligence and a machine learning algorithm have been used to provide a new direction for the classification of kidney diseases. To the best of our knowledge, our study is the first to combine artificial intelligence and urinary proteomic mass spectra information to the diagnosis and classification of kidney diseases.

Compared with RF, SVM, and ANNs, the XGBoost model with mass spectra information for urinary proteomics has shown a perfect performance for the diagnosis of kidney disease. This is consistent with the classification ability of XGBoost models when applied to other clinical diseases. Therefore, compared with other machine learning algorithms, the advantages of the XGBoost algorithm are as follows^27^: First, XGBoost adds a regularization term to the objective function, which reduces the variance of the model, simplifying the model while preventing an over fitting. Second, XGBoost not only uses the first derivative, it also uses the second derivative to make the loss more accurate. Third, when the training data are sparse, the default direction of the branch can be specified for a missing or specified value, which can significantly improve the efficiency of XGBoost. Fourth, XGBoost supports column sampling and parallel optimization, thereby reducing the number of computations and improving the efficiency. The peak value of the urine proteome mass spectrometry data is presented in the form of a set of numbers in abscissa and ordinate coordinates, which is used in the construction of the XGBoost model to provide full play to such advantages.

In our study, the overall accuracy of the diagnostic XGBoost model for the four groups was 96.03%, which is basically consistent with the accuracy of a renal biopsy. Therefore, it highlights the advantages brought about by a non-invasive diagnostic method used for an artificial intelligence model applied to proteomics. In addition, we conducted a detailed assessment of the modelling accuracy for each type of kidney disease and the HC. The specificity of the diagnosis model for the four types was more than 95% (97.07%–99.91%), and thus its misdiagnosis rate is extremely low and its ability to distinguish each type of disease shows excellent stability. Although the sensitivity of the four types was approximately 90% (89.81%–97.63%), the sensitivity of the three types of kidney disease, excluding the HC group, were lower than that of the specificity. Therefore, the missed diagnosis rate of this model is higher than the misdiagnosis rate, which indicates that this model may be more suitable for an accurate disease diagnosis than for disease screening. The next steps of this research will focus on an improvement of the prediction sensitivity of the model. For all four types, the sensitivity of this model regarding a DKD diagnosis is the worst (89.81%), the reason for which may be the smaller number of DKD patients included, the smaller average number of spectra, or the significant differences with the other groups. Among them, the low average number of urinary proteomic mass spectrum analyses is a response to the state of a real disease, which cannot be avoided. The authors hope that more samples will be included in the next study to reduce the problems caused by a data imbalance. In addition, through an analysis of the ROC curve, gain plot, and Lorenz curve, this study showed that the model achieves strong robustness and a high accuracy.

At present, a few existing XGBoost models for kidney disease diagnosis have been constructed using data on the clinical characteristics and individual laboratory test indicators. Ogunleye et al.^7^ applied 250 CKD cases and 150 HC groups to train and validate the XGBoost model with 22 clinical features. The accuracy, sensitivity, and specificity of this XGBoost model were all 100%. Xiao et al.^23^ also constructed a XGBoost model for the prediction of CKD progression, including 551 patients with proteinuria. A total of 13 blood-derived tests and 5 demographic features were used as variables to train the model. The accuracy of this progression model was 0.87. Applying 36 characteristics of 2,047 Chinese patients from 18 renal centers, Chen et al.^28^ used a XGBoost model for a prediction of the end-stage CKD. The C statistic of this XGBoost model was 0.84. Because all of these reports were constructed using clinical information and the outcome indicators were inconsistent, poor comparability with our diagnostic XGBoost model was achieved. However, the accuracy of our model is high.

The KDClassifier classified the characteristic differences for different pathological types of CKD at the level of integrated information of mass spectrometry proteomics found in urine. There are no specific proteins or laboratory indicators of clinical concern, such as GFR, urine protein, or creatinine. This is significantly different from our normal assumption. The information captured by a machine learning model is more abundant than the comparative analysis of differential proteins. How to explain the specific content of the information captured by machine learning with human logic requires further research and discussion.

Overall, the KDClassifier, an XGboost diagnostic model, established in this study showed its feasibility and superiority for clinical application. However, in terms of economics, the cost of a mass spectrometry analysis of the proteomics is at present relatively high, and there is still a long way to go regarding its clinical application. With further innovations in science and technology, however, we expect the cost of mass spectrometry analysis to inevitably decline. The KDClassifier is not only suitable for the classification of the three types of kidney disease considered, it also has the potential to be extended to all types of kidney disease. The diagnostic advantages of this model will be fully demonstrated.

Our study also has certain limitations. First, the cohort used is not from a prospective trial, and selective bias is inevitable. Second, only three common types of kidney disease were included. Whether this learning machine diagnostic method is suitable for other types of kidney diseases needs further research and validation using a larger sample size. Third, owing to a relatively small sample size, we did not include more clinical parameters for an AI-assisted analysis. If we include more clinical data, it will further improve the diagnostic power. Fourth, this study only attempted to compare four mainstream machine learning methods with certain limitations. Fifth, only the mass spectra of urinary proteomic information was used, and the clinical information of the patients was omitted. If both types of information are combined, the patients can be better diagnosed. We expect to develop more suitable artificial intelligence algorithms for a noninvasive and accurate diagnosis of kidney diseases.

## CONCLUSION

In conclusion, KDClassifier, a machine learning classification model that applies information on mass spectra from urinary proteomics showed a high accuracy in the diagnosis of different types of CKD. This study provided a new idea for applying artificial intelligence in the accurate and non-invasive diagnosis of kidney diseases. In addition, KDClassifier provides a potential tool for the classification of all types of kidney diseases.

## METHODS

### Study Population

In this study, a total of 202 urine samples from IgAN (n = 50), MN (n = 50), and DKD (n = 34) patients and from a healthy control (HC) group (n = 68) were collected in tubes in accordance with standard hospital operating procedures. All patients with kidney disease were examined through a renal biopsy, and secondary types of IgAN or MN were excluded. The urine samples were collected within 1 week before the renal biopsy. Briefly, the midstream urine from the second morning void was collected in appropriate containers and centrifuged at 1,000 × g for 20 min. The precipitate was discarded, and 500 μL of the supernatant (including the soluble proteins, exosomes, and other membrane elements) was collected in a 1.5-mL tube and stored at –80 °C until use. Diagnosis and pathological examinations of the kidney diseases were conducted at the Department of Nephrology, Sichuan Provincial People’s Hospital. Informed consent was obtained from the patients. The study protocol was approved by the Medical Ethics Committee of the Sichuan Provincial People’s Hospital and West China Hospital.

### Urinary Protein Digestion

Human urinary protein digestion was conducted using a filter-aided sample preparation. Each 100-μL urine supernatant was loaded onto a 30-kDa ultrafiltration device. After centrifuging at 13,000 × g for 15 min, a 100-μL UA solution with 20 mM DTT was added and reacted for 4 h at 37 °C. An alkylation reaction was then achieved by adding a 100-μL UA solution with 50 mM iodoacetamide (IAA) and incubated in the dark for 1 h at room temperature. The buffer was replaced with 50 mM ammonium bicarbonate. Finally, 10 μg trypsin was added to each filter tube and the reaction was maintained for 16 h at 37 °C. The digestion was collected, and the concentrations were measured at 480 nm. The urinary protein digestions were freeze-dried and stored at −80 °C.

### Mass Spectrometer Analysis

A urinary peptide analysis was conducted using an Orbitrap Fusion Lumos mass spectrometer (Thermo Fisher Scientific, Waltham, MA, USA). Briefly, the peptides were dissolved in 0.1% FA and separated into a 75-μm I.D. column 15 cm in length over a 78-min gradient (buffer A, 0.1% FA in water; buffer B, 0.1% FA in 80% ACN) at a flow rate of 300 nL min^-1^. MS1 was analyzed with a scan mass range of 300–1,400 at a resolution of 120,000 at 200 m/z. The RF lens, automatic gain control (AGC), maximum injection time (MIT), and exclusion duration were set at 30%, 5.0 e^5^, 50 ms, and 18 s, respectively. MS2 was analyzed in data-dependent mode for the 20 most intense ions. The isolation window (m/z), collision energy, AGC, and MIT were set at 1.6, 35%, 5.0 e^3^, and 35 ms, respectively.

### Spectra Establishment

The RAW data from a mass spectrometer were converted into MGF format files, with each file containing thousands of pieces of mass spectrum information. The x-coordinate is the mass-to-charge ratio (m/z), and the y-coordinate is the relative peak intensity. The mass spectra from each file are used to profile the urinary proteome of each patient. The data on the mass spectrometry from the urinary proteomics were deposited at the ProteomeXchange Consortium through the PRIDE partner repository with the dataset identifier PXD018996.

### Data Pre-processing

These mass spectra contain four classes of CKD urinary proteomic information (IgAN, MN, DKD, and HC). The MGF format files were processed using an illumination normalization method. The data of all original urinary proteomic mass spectra were transformed into double column arrays of indefinite length (with the abscissa and ordinate values of the peaks in the spectrum). Owing to the unequal length in the different arrays, we set an array with a length of 50 rows (the maximum value). We then merged every double column array into a single feature data line with a length of 100. Data of insufficient length were considered as missing values. Finally, a dataset with four different data labels (IgAN, MN, DKD, and HC) were built and imported into the XGBoost model.

### XGBoost Model

XGBoost is a machine learning technique developed by Chen et al. that assembles weak prediction models^25^. It generates a series of decision trees in a gradient boosting manner, which means that it generates the next decision tree based on the current tree to better predict the outcome. After training, a classification prediction system composed of a series of decision trees is achieved. This is an extendible and cutting-edge application of a gradient boosting machine and has been proven to push the limits of computing power for boosted tree algorithms. Gradient boosting is an algorithm in which new models are created for predicting the residuals of the prior models, and then added together to make the final prediction. A gradient descent algorithm is used to minimize the loss when adding new models. XGBoost with a multi-core CPU reduces the look up times of the individual trees created. With this algorithm, the definition of the *K* additive function ensemble model (K trees) is given as follows:

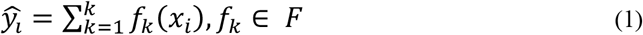

where *x*_*i*_ indicates the *i*^*th*^ sample, *F* is the space containing all trees, and *f*_*k*_ refers to the *k*^*th*^ function in functional space *F*.

To train the ensemble model, the objective is minimized as follows:

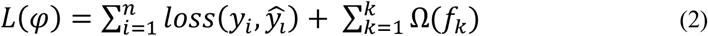

Here, *loss* is a loss function that measures the difference between the target *y*_*i*_ and prediction *ŷ*_*l*_. In addition, Ω penalizes the complexity and is defined as

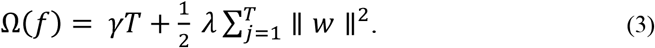

The number of leaves in a tree is defined as *T*; in addition, *γ*indicates the minimum loss reduction, *λ* is the weight of the regularization, and ∥ *w* ∥ represents the corresponding score of the leaves.

The XGBoost algorithm can handle missing data automatically by adding a default direction for the missing values in each tree node. The default direction is learned during the training procedure. When a value is missing in the validation data, the instance is classified into the default direction. This means that we only need to input a reduced number of important variables while leaving the others as null values during the application stage.

We maintained 20% of the data as the validation set and the remaining 80% to train our diagnosis XGBoost model. The hyperparameters used in our analysis were as follows: the learning rate = 0.01, the minimum loss reduction = 10, the maximum tree depth = 10, the number of subsample = 0.8, the number of trees = 300, and the number of rounds = 100. A simultaneous grid search over gamma, reg lambda, and the subsample was used to re-examine the model and check for differences between the optimum values.

### Other Machine Learning Models

Random forest (RF) is a type of classifier that uses randomly generated samples from existing situations and consists of multiples trees^29^. To classify a sample, each tree in the forest is given an input vector, and a result is produced for each tree, and the tree with the most votes is chosen as the result. RF divides each node into branches using the best randomly selected variables on each node.

An SVM is a controlled classification algorithm based on statistical learning theory^30^. The working principle of an SVM is based on the principle of predicting the most appropriate decision function that separates the two classes; in other words, based on the definition of a hyperplane, it can distinguish two classes from each other in the most appropriate manner possible. Similar to a classification, kernel functions are used to process nonlinear states during the regression. In cases in which the data cannot be separated linearly, non-linear classifiers can be used instead of linear classifiers. An SVM transforms into a high dimensional feature space, which can be easily classified linearly from the original input space by means of a nonlinear mapping function. Thus, instead of finding values by repeatedly multiplying them using kernel functions, the value is directly substituted in the kernel function, and its counterpart is found in the feature space. In this way, there is no need to deal with a space with a very high-dimensional quality. An SVM has four widely used kernel functions, namely, linear, polynomial, sigmoid, and radial basis functions.

Artificial Neural Networks (ANNs) make up an information processing system inspired by biological neural networks and includes some performance characteristics similar to those of biological neural networks^31^. The simplest artificial neuron consists of five main components: inputs, weights, transfer function, activation function, and output. In an ANN, neurons are organized in layers. The layer between the input and output layers is called the hidden layer. The network is regulated by minimizing the error function. The connection weights are re-calculated and updated to minimize the error. Thus, it is aimed at bringing output values that are closest to the ground truth values of the network.

### Performance Evaluation and Statistical Analysis

We divided all mass spectrum data from the CKD urinary proteomics into a training dataset (80%) and a validation dataset (20%). The training dataset was directly used to train the framework and create a diagnosis model using XGBoost, RF, an SVM, and an ANN. The validation dataset was used to calculate the diagnostic accuracy. We compared the accuracy of the four machine learning models, and constructed a confusion matrix to calculate the sensitivity, specificity, positive predictive value, and negative predictive value of the XGBoost diagnosis model.

We also constructed ROC curves for the CKD diagnosis model. We calculated the AUC of the ROC curves to evaluate the prediction capabilities of the diagnosis model. Lorenz and gain curves were then constructed to evaluate the goodness of fit of the XGBoost diagnosis model.

The Lorenz and gain curves were established as graphical representations of the distribution of the econometrics, and have been proven to be valuable analytic tools in other fields as well, including in the evaluation of classifier models. Kendall and Stuart introduced a Lorenz curve arranged in ascending order according to the probability returned by the classification model. Dividing 0-1 equally into N parts, the divided points are the threshold (abscissa), and the true positive rate (TPR) and false positive rate (FPR) are calculated. Taking the TPR and FPR as ordinates, draw two curves, i.e., Lorenz curves (or KS curves). The cut-off point (KS value) is the position where the distance between the TPR and FPR curves is the largest. A KS value of more than 0.2 is considered a good prediction accuracy. The gain plot is an index used to describe the overall accuracy of the classifier models. With an increase in the depth, the gain rate of the classifier model is compared with the natural random classification model. The steeper the curve and the larger the slope, the better the TPR obtained by the model.

Continuous variables are expressed as the mean ± standard deviation and are compared using a T test. Categorical variables are expressed as percentages, and a Chi-square test or Fisher’s exact test was employed to compare the differences in the variables. SPSS software version 22.0 (IBM Corp) was used for a comparative analysis of the basic characteristics. The machine learning models were developed using Python 3.4.3 (using the XGBoost library, DF library, SVM library, and ANN library). The evaluation and analysis method for determining the performance of the XGBoost model (KDClassifier) applied R (using the pROC, dplyr, caret, lattice, and ggplot2 packages) version 3.5.2. The 95% confidence intervals (CIs) were then calculated. All *P* values were two tailed, and a *P* of less than 0.05 was considered statistically significant.

## Data Availability

The mass spectra data on the urinary proteomics were deposited into the ProteomeXchange Consortium through the PRIDE partner repository using the dataset identifier PXD018996.

## ACKNOWLEDGMENTS

This work was funded by grants from the National Natural Science Foundation of China (grant no. 31901038), the China Postdoctoral Science Foundation (2019M653438), the Post-Doctoral Research Foundation of West China Hospital of Sichuan University (2018HXBH062), and the 1.3.5 Project for Disciplines of Excellence, West China Hospital, Sichuan University (ZYGD18014, CJQ).

